# Psilocybin-induced reduction in chronic cluster headache attack frequency correlates with changes in hypothalamic functional connectivity

**DOI:** 10.1101/2022.07.10.22277414

**Authors:** Martin K. Madsen, Anja Sofie Petersen, Dea S. Stenbæk, Inger Marie Sørensen, Harald Schiønning, Tobias Fjeld, Charlotte H. Nykjær, Sara Marie Ulv Larsen, Maria Grzywacz, Tobias Mathiesen, Ida L. Klausen, Oliver Overgaard-Hansen, Kristoffer Brendstrup-Brix, Kristian Linnet, Sys S. Johansen, Patrick M. Fisher, Rigmor H. Jensen, Gitte M. Knudsen

## Abstract

Chronic cluster headache (CCH) is an excruciating disorder of unknown pathophysiology, but hypothalamic dysfunction has been implicated. CCH is difficult to treat but on a case-basis, the psychedelic compound psilocybin is said to have beneficial effects. In this first-ever clinical trial (NCT04280055), we evaluate in a small open-label study of CCH patients the feasibility and prophylactic effect of three low-to-moderate doses of psilocybin as well as effects on hypothalamic functional connectivity (FC), using functional magnetic resonance imaging. The treatment was well-tolerated and without serious adverse reactions. Attack frequency was on average reduced by 30% from baseline to follow-up (P_FWER_=0.008). One patient experienced 21 weeks of complete remission. Changes in hypothalamic-diencephalic FC correlated negatively with relative reduction in attack frequency, implicating this neural pathway in treatment response. Further clinical studies are warranted to confirm the safety and prophylactic efficacy of psilocybin for CCH and hypothalamic involvement in treatment response.

## Main text

Cluster headache (CH) is one of the most painful conditions known and affects 0.1% of the population, a prevalence similar to Parkinson’s disease.^1^ CH exists in two forms: episodic or chronic (CCH).^2^ Ten to fifteen percent of patients with CH have CCH, which means less than three months without attacks during a year.^2^ CH is frequently associated with anxiety, depressive symptoms and suicidal ideation.^3^ Medical treatment consists of acute abortive treatment and prophylactic treatment. Although the standard prophylactic options (e.g., verapamil or lithium) substantially reduces CH attack frequency in many patients, the treatment may have unacceptable side effects or insufficient treatment response. Thus, novel treatments are needed.

Observational reports indicate that the psychoactive constituent of *magic mushrooms*, psilocybin, and other serotonin 2A receptor (5-HT2AR) agonists may have beneficial prophylactic effects.^4–6^ As of today, the potential prophylactic effect of psilocybin on CH has not been formally investigated in a prospective clinical trial.

Here we evaluate the feasibility and prophylactic effects of psilocybin in patients with CCH in a small open-label study. Since hypothalamic dysfunction is believed to play a key role in CH pathophysiology,^7–10^ we also assess the association of treatment response with hypothalamic functional connectivity (FC), measured using functional magnetic resonance imaging (fMRI).

Ten patients (5 females, mean age (SD) = 49.4 (12.9)) with a verified diagnosis of CCH completed the study. A moderate dose of peroral psilocybin (0.14 mg psilocybin per kg body weight) was administered three times, spaced by seven days. The psilocybin used was COMP360, COMPASS Pathways’ proprietary pharmaceutical-grade synthetic psilocybin formulation that has been optimized for stability and purity. The patients abstained from regular prophylactic therapies and recorded daily the number of headache attacks and average pain intensity (one combined estimate for each day, numerical rank scale: 0-10) during the study period (four-week baseline, two-week treatment period, and four-week follow-up) (**Table 1, Fig 1 a & b**). The primary outcomes were assessment of feasibility and safety, change in headache attack frequency from baseline to follow-up, and effects on hypothalamic functional connectivity (FC). Patients were contacted three and six months after the ten-week study period to probe the duration of potential remission and patients’ view of the treatment.

**Table 1.**
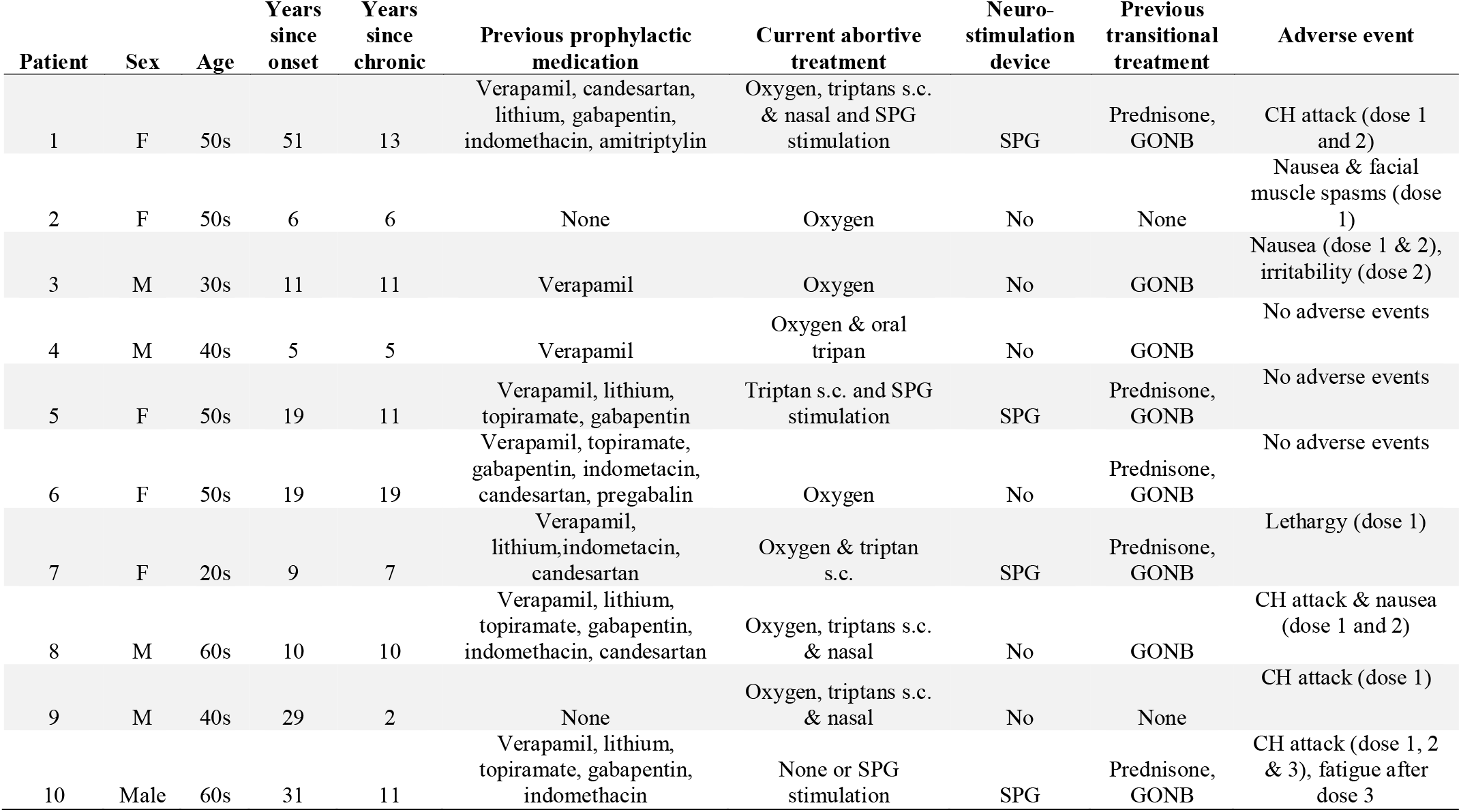
Characteristics of study patients and reported adverse events. F: female, M: male. SPG: sphenopalatine ganglion stimulation device, GONB: greater occipital nerve block, s.c.: subcutaneous.

| Patient | Sex | Age | Years since onset | Years since chronic | Previous prophylactic medication | Current abortive treatment | Neuro-stimulation device | Previous transitional treatment | Adverse event |
| --- | --- | --- | --- | --- | --- | --- | --- | --- | --- |
| 1 | F | 50s | 51 | 13 | Verapamil, candesartan, lithium, gabapentin, indomethacin, amitriptylin | Oxygen, triptans s.c. & nasal and SPG stimulation | SPG | Prednisone, GONB | CH attack (dose 1 and 2) |
| 2 | F | 50s | 6 | 6 | None | Oxygen | No | None | Nausea & facial muscle spasms (dose 1) |
| 3 | M | 30s | 11 | 11 | Verapamil | Oxygen | No | GONB | Nausea (dose 1 & 2), irritability (dose 2) |
| 4 | M | 40s | 5 | 5 | Verapamil | Oxygen & oral tripan | No | GONB | No adverse events |
| 5 | F | 50s | 19 | 11 | Verapamil, lithium, topiramate, gabapentin | Triptan s.c. and SPG stimulation | SPG | Prednisone, GONB | No adverse events |
| 6 | F | 50s | 19 | 19 | Verapamil, topiramate, gabapentin, indometacin, candesartan, pregabalin | Oxygen | No | Prednisone, GONB | No adverse events |
| 7 | F | 20s | 9 | 7 | Verapamil, lithium, indometacin, candesartan | Oxygen & triptan s.c. | SPG | Prednisone, GONB | Lethargy (dose 1) |
| 8 | M | 60s | 10 | 10 | Verapamil, lithium, topiramate, gabapentin, indomethacin, candesartan | Oxygen, triptans s.c. & nasal | No | GONB | CH attack & nausea (dose 1 and 2) |
| 9 | M | 40s | 29 | 2 | None | Oxygen, triptans s.c. & nasal | No | None | CH attack (dose 1) |
| 10 | Male | 60s | 31 | 11 | Verapamil, lithium, topiramate, gabapentin, indomethacin | None or SPG stimulation | SPG | Prednisone, GONB | CH attack (dose 1, 2 & 3), fatigue after dose 3 |

**Fig 1.**
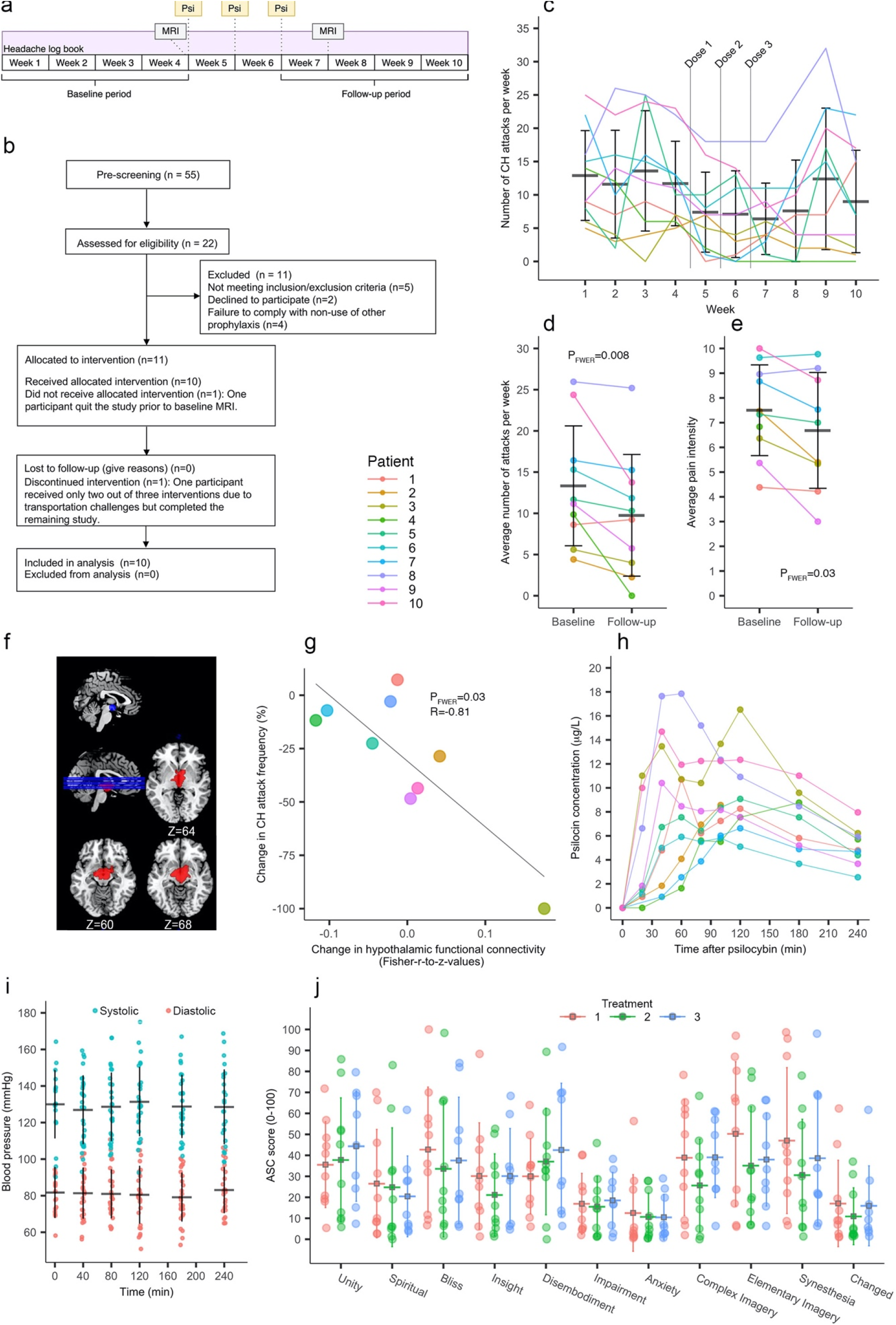
Effects of psilocybin in patients with chronic cluster headache. **a**) Study design. MRI: magnetic resonance imaging. Psi: psilocybin treatment. MRI data acquisition was done the day before the first psilocybin treatment and one week after the last treatment. Plasma psilocin level (PPL) was measured during the first treatment; psychoactive effects and blood pressure were measured on all treatment days. **b**) Flow-chart of study. **c**) Attacks per week. CH: cluster headache. **d**) Effect of psilocybin on weekly attack frequency. Statistical test: Wilcoxon signed-rank test. **e**) Effect of psilocybin on average attack pain intensity. **f**) Hypothalamic 8 mm radius sphere seed (blue) centered on posterior hypothalamus and diencephalic cluster (red) exhibiting significant functional connectivity with the seed region across baseline and follow-up scans. **g**) Negative correlation between percent change in attack frequency and change in functional connectivity between seed region and diencephalic cluster. Statistical test: simple linear regression. **h**) Individual subject PPL measured at the first treatment. It was not possible to obtain blood samples after the 100 min measurement for Patient 2 due to IV access failure. **i**) Blood pressure measurements pooled across all three treatments. **j**) Psychometric evaluation of psychoactive effects. Scores on the 11-dimension Altered States of Consciousness (ASC) questionnaire rated at the end of each treatment day. Error bars show mean and standard deviation.

First, we evaluated the feasibility and safety of performing the psilocybin treatments. Nine patients completed all three psilocybin treatments; one patient was due to logistical issues unable to attend the third treatment but completed the rest of the study. The dosing regime was well tolerated by all patients, and we observed no serious adverse reactions (**Table 1, SI Table 1)**. At least one patient (Patient 5) reported beneficial long-lasting psychological effects, lasting at least six months (**SI Table 1**). Psilocybin induced variable acute subjective psychoactive effects (**Fig. 1j**) but was not associated with changes in blood pressure (**Fig. 1i**). Interestingly, one patient (Patient 6) reported complete absence of psychoactive effects of psilocybin; objective effects (e.g., dilated pupils) were also imperceptible to the staff. This patient also had the lowest peak concentration (C_max_) of plasma psilocin (PPL) of all patients (5.9 µg/L).

We then examined the prophylactic effects of psilocybin by comparing the four-week baseline to the four-week follow-up. We observed a statistically significant reduction in headache frequency, (P_FWER_ = 0.008, mean change (SD) = -3.57 (3.87) attacks/week, percent change (SD): -30 (31)% (**Fig. 1 c, d**)). Notably, one patient (Patient 4) experienced complete remission for 21 weeks, which began one day after the first psilocybin session. Effects of psilocybin treatment on average self-rated pain intensity showed a statistically significant reduction from baseline to follow-up (P_FWER_ = 0.03, mean change (SD) = - 0.89 (0.94) attacks/week, percent change (SD): -13 (15)% (**Fig. 1e)**). Further, the patients used acute therapy for their attacks at the overall same rate during follow-up and baseline (**SI Fig 1**). Self-reported average duration of all CH attacks occurring each day was unchanged (**SI Fig. 2)**.

After establishing prophylactic effects, we investigated effects on hypothalamic FC. Due to a Covid-19-related lock-down, MRI data was incomplete for one participant, giving a total sample size of nine patients for the FC analysis. Four clusters showed significant positive FC with an 8 mm radius seed centered on the posterior hypothalamic, across baseline and follow-up scans: 1) a large diencephalic cluster encompassing the hypothalamus, thalamus, caudate and brain, 2) a cluster in the left lingual gyrus, 3) a cluster in the right cerebellum, and 4) a) cluster in occipito-temporal white matter (**SI Table 3**). Linear regression analysis showed a significant negative correlation between percent change in CH attack frequency and change in hypothalamic FC with the diencephalic cluster (p_FWER_=0.03, R=-0.81, **Fig. 1g**). We found no significant associations for the remaining three FC estimates (P-values > 0.7), and no significant change in hypothalamic FC from baseline to follow-up (P-values > 0.6). Exploratory analysis of effects on networks FC and on brain structure is available in **SI** (**SI Table 2, SI Fig 3**).

We observed substantial interindividual variability in PPL (**Fig. 1h**): maximum PPL concentration (C_max)_) (mean (SD) = 11.8 (4.3) μg/L, median[range] = 10.4[5.9;17.9] μg/L and area under curve (AUC) (mean (SD) 1711 (682) μg*min/L, median[range] = 1483[968;2615] μg*min/L. PPL from one patient was not included due to missing data (IV access failure). Exploratory linear regression analyses did not show significant associations between PPL C_max_ or AUC with change in attack frequency or pain intensity (all P-values > 0.9).

Here, we provide the first formal clinical evidence for beneficial prophylactic psilocybin effects in CCH. The treatment was safe, well-tolerated and was associated with a substantial but variable reduction in CH attack frequency, supporting that psilocybin therapy may constitute a valuable therapeutic option in some patients with CCH.

Interestingly, we observed a negative correlation between percent reduction in attack frequency and FC changes between the posterior hypothalamus and a large diencephalic cluster (**Fig. 1 f & g**). Hypothalamic dysfunction is a prime candidate for pathophysiological involvement in CH, as supported by several neuroimaging studies.^7–10,12^ Psilocybin acutely alters the brain’s normal functional architecture, including reduced functional integrity of individual functional networks, increased FC between networks, and increased thalamic FC.^13^

Accumulating evidence suggest increased neuroplasticity after psilocybin.^14,15^ We speculate that a recalibration of aberrant hypothalamic-diencephalic neuronal pathways, supported by increased neuroplasticity, may underlie at least part of psilocybin’s prophylactic potential in CH.

Previous psilocybin CH survey studies do not specifically report on reduced pain intensity, making it difficult to directly compare our results.^4,5,16,17^ We observed a 13% reduction in pain intensity. Given that the study is an open-label design, we cannot exclude that at least a part of the observed pain-reducing effect is presumably due placebo, the true effect size is most likely small.

Interestingly, one patient reported no subjective psychoactive effects of psilocybin. Although this patient displayed the lowest level of PPL, we have previously observed notable psychoactive effects at this PPL in healthy individuals.^13,18^ Interestingly, the patient displayed a reduction in headache frequency of 22% from baseline to the follow-up, indicating a possible prophylactic effect even in the absence of psychoactive effects.

One patient reported enhanced quality of life and greater appreciation for life despite long-term unchanged headache burden. This is consistent with reported long-lasting positive psychological effects in both patients and healthy individuals.^19^ It is possible that psilocybin treatment could be a valuable psychological intervention in patients with CCH or other chronic pain conditions, an effect that may be linked to psilocybin-induced increased mindfulness as we have previously observed in healthy individuals.^20^

Although psilocybin overall has a favorable safety profile,^21^ it is important to emphasize that psilocybin has potent psychedelic properties, is illegal in most countries, and that serious negative health-related outcomes have been observed in non-clinical settings due to poorly regulated behavior.^22^ Thus, we caution that our results are preliminary and advise that patients abstain from self-medicating with psychedelics.

Our study is not without limitations. Our original aim of including 20 patients was hampered by covid-19-related societal and institutional restrictions and data from ten patients only were available for data analysis, and we recognize the small sample size limits the ability to draw strong statistical inference and to observe rare side-effects. Further, we did not employ a placebo-control condition. The included patients were relatively treatment refractory to standard prophylactic treatment, and recruited from a tertiary headache center, which may limit the generalizability to other CH-patients. Intake of prophylactic medication was not allowed in our trial, and it is also possible that our results do not generalize to patients on regular prophylactic agents.

In conclusion, the administration of three moderate doses of psilocybin, conducted in a controlled clinical setting, was feasible and the treatment safe and tolerated well by all patients. Both attack frequency and pain intensity decreased after the treatments. Changes in hypothalamic-diencephalic FC correlated negatively with percent change in attack frequency, implicating a neural pathway in treatment response. Although encouraging, future placebo-controlled studies in carefully controlled settings are needed to confirm the clinical utility of psilocybin in CH prophylaxis.

## Data Availability

All data produced in the present study are available upon reasonable request to the authors

## Acknowledgements

The assistance of Drummond McCulloch, Lone Freyr, Arafat Nasser, Annette Johansen, Dorthe Givard, Peter Jensen, Kristian Jensen, and Martin Snoer is gratefully acknowledged. COMPASS Pathways Ltd, UK, kindly provided the study drug. The study was sponsored by Augustinusfonden (grant number: 18-0317), Rigshospitalet’s Research Council (grant numbers: R130-A5324 and R214-A9447), Innovation Fund Denmark (NeuroPharm, grant number: 4108-00004B), and Lundbeck Foundation through Danish Neurological Society (no grant number available).

## Contributions

Conceptualization: MKM, GMK, RHJ, ASP, DSS. Grant applications: MKM, GMK, RHJ. Data collection: MKM, ASP, DSS, IMS, HS, TF, CHN, SMUL, MG, TM, TLK, OOH, KBB. Psilocin analysis: KL, SSJ. Data analysis: MKM, PMF, GMK, PMF. Writing original draft: MKM, GMK. All authors have engaged in interpretation of the data, critical review of the present manuscript and have approved of and are responsible for the final version.

## Ethics declarations

The study was approved by the ethics committee for the capital region of Copenhagen, Denmark (H-18040896 with amendments 64708, 65371, 65944, 71621, 72461) and the Danish Medicines Agency (EudraCT 2018-003382-34, amendment 20191209).

## Data availability

Data can be made available upon reasonable request, as permitted by applicable law.

## Methods and Materials Patients

Ten patients (5 females, mean age (SD) = 49.4 (12.9)) with a verified diagnosis of CCH were recruited from Danish Headache Center, Glostrup Hospital, Denmark (**Table 1**) and completed the study. The study was approved by the ethics committee for the capital region of Copenhagen (journal identifier: H-18040896, H-amendments: 64708, 65371, 65944, 71621, 72461) and Danish Medicines Agency (EudraCT identifier: 2018-003382-34, amendments: 20191209), ClinicalTrials.gov identifier: NCT04280055.

Inclusion criteria were: 1) age 18-65 years, 2) a diagnosis of CCH according to IHCD-III,^2^ 3) ability to separate cluster headache attacks from other types of headache, 4) a history of at least 4 attacks per week in the last 4 weeks before inclusion. Exclusion criteria were: 1) a history of using a serotonergic hallucinogen for CH, 2) participation in any clinical trials within 30 days preceding study enrollment, 3) use of other prophylactic CH medication within the last two weeks, 4) current use of drugs suspected to interfere with treatment (e.g. antipsychotic medication) or to be hazardous in combination with psilocybin, 5) presence of other trigeminal autonomic cephalalgias than CH, 6) known hypersensitivity or allergy to multiple drugs (including psilocybin), 7) a history of or current medical or psychiatric condition that might render participation unsuitable, 8) present or previous manic or psychotic disorder or critical psychiatric disorder, 9) current drug or alcohol abuse, 10) MRI contraindications, 11) pregnancy or breastfeeding, 12) not using safe contraception (if fertile woman), 13) stroke (<1 year from inclusion), 14) myocardial infarction (<1 year from inclusion), 15) hypertension (> 140/90 mmHg at inclusion), 16) clinically significant cardiac arrhythmia (<1 year from inclusion).

All patients were thoroughly informed about the study, including possible effects and side-effects of psilocybin. After giving informed consent, all patients underwent a medical examination, including neurological and somatic examination, blood screening panel for common biomarkers, ECG, and a screening for present and past psychiatric disorders using the Mini-International Neuropsychiatric Interview, Danish Translation, version 6.0.0.^23^ Eleven patients were included in the study (**Fig 1b**).

### Procedures

After study inclusion, patients completed an online headache logbook daily for ten weeks. The information entered in the headache logbook included the number of CH attacks and estimated average pain intensity (i.e., one summary estimate for all attacks prior to acute treatment (0-10, numeric rank scale) on that day), and the number of times of acute treatment use (either oxygen or sumatriptan s.c.). A four-week baseline period was followed by three psilocybin treatments spaced by one week, and then followed by a four-week follow-up period (for study design, see **Fig 1a**). The day prior to the first psilocybin treatment and one week after the last treatment, patients underwent an MRI scan session. Patients were contacted by phone after three and six months to obtain information of potential remission duration and perception of the psilocybin treatment (**SI Table 1**).

### Psilocybin treatments

On the day before the first psilocybin treatment, every participant met with two trained session facilitators, who also provided psychosocial support on the treatment day, following an established protocol at our lab previously employed in healthy participants.^13,18,20^ Psilocybin dose was determined according to body-weight at inclusion (0.14 mg psilocybin per kg bodyweight), using 1 mg and 5 mg capsules/tablets (COMP360, COMPASS Pathways’ proprietary pharmaceutical-grade synthetic psilocybin formulation that has been optimized for stability and purity). The 0.14 mg/kg dose and pulse treatment regimen were determined based on patient accounts that low to moderate psilocybin doses spaced by 4-7 days may be effective.^24^ Dose was also determined based on our previous psilocybin 5-HT2AR positron emission tomography (PET) occupancy study, indicating that 0.14 mg/kg produces high psilocin occupancy at cerebral 5-HT2ARs while still having limited psychoactive effects.^18^ Patients were asked to limit to a minimum the intake of food and caffeine on the morning of all treatment days and were asked to standardize their food intake across all three treatment days. On the first treatment day, blood samples were acquired before and after psilocybin administration for the purpose of analyzing plasma psilocin level (PPL). At the end of treatment days, when patients and staff assessed that the psychoactive effects had waned sufficiently, patients completed the 11-dimension (11-D) Altered States of Consciousness (ASC) questionnaire, which quantifies eleven dimensions of subjective experience during psilocybin.^25,26^ After each psilocybin treatment, as per our standard, patients met the day after with the attending session facilitators, which allowed for patients to share and integrate their experience.

### Blood pressure measurements

Blood pressure was measured immediately prior to psilocybin administration and at 40, 80, 120, 180, and 240 minutes after drug administration, using an electronic blood pressure measurement device (Omron M3 Comfort, Kyoto, Japan).

### Plasma psilocin concentration

Blood samples were drawn from an intravenous catheter placed in the antecubital vein; sampling was done immediately prior to psilocybin intake and 20, 40, 60, 80, 100, 120, 140, and 240 minutes after. Blood samples were always obtained after blood pressure measurement. PPL was determined using ultra high performance liquid chromatography and tandem mass spectrometry, as previously described^18^ with minor changes. The transition of the internal standard psilocin-d10 was m/*z* 215 -> 65.8 with collision energy of 14 eV corresponding to the quantitative transition of psilocin. The reconstitution mixture after precipitation and evaporation were added 1 mM ascorbic acid.

### Magnetic resonance imaging

Structural and functional MRI data was acquired using a 32-channel head coil on a 3T Siemens Prisma scanner (Siemens, Erlangen, Germany). Blood oxygen level dependent (BOLD) fMRI data was obtained using a T2*-weighted gradient echo-planar imaging (EPI) sequence (TR = 800 ms, TE = 37 ms, flip angle = 52°, in-plane resolution=2×2×2 mm, 72 slices (thickness = 2.0 mm, no gap), multiband acceleration factor = 8). For each BOLD fMRI data acquisition, 375 volumes were acquired (5 min) for the first five patients and for the last five patients, 750 volumes were acquired (10 min). Participant were instructed to keep their eyes closed, let their mind wander freely and not fall asleep. A high-resolution, T1-weighted 3D structural MP-RAGE image was also acquired during each scan session (inversion time = 920 ms, TE = 2.41 ms, TR = 1810 ms, flip angle = 9°, resolution = 0.8×0.8×0.8 mm, 224 slices; slice thickness = 0.8 mm, no gap). Nine individuals completed both baseline and follow-up scan sessions; a covid-19-related lock-down prevented acquisition of follow-up MRI data in one patient.

### Image preprocessing

BOLD image preprocessing was performed in SPM12 (http://www.fil.ion.ucl.ac.uk/spm), including unwarping, realignment, co-registration, segmentation of structural image and normalization to Montreal Neurological Institute (MNI) space.

Structural images underwent cortical reconstruction and volumetric segmentation using FreeSurfer v7.1 (http://surfer.nmr.mgh.harvard.edu), and all images were visually inspected for white and pial surface estimations and manually corrected accordingly. Fifteen regions as defined by the Desikan-Killiany atlas^27^ were selected for structural regional analysis. For each region, average cortical thickness and grey matter (GM) volume were extracted. Hippocampal volumes as well as both cortical and total GM volumes were extracted from the volumetric segmentation.

### Functional connectivity analysis

Denoising, which included band-pass filtering, motion and spike regression and principal-component based noise-reduction (aCompCor),^28^ and FC estimation was performed using CONN (v17c) as previously described.^13^ FC (Fisher-transformed R-to-z values) between an 8 mm radius sphere seed placed in the posterior hypothalamus (MNI coordinates: 3, -9, -10)^29^ and the rest of the brain was conducted across all images using both baseline and follow-up BOLD images (statistical significance threshold: voxel-level p-value threshold = 0.001, cluster-level significance level p_FWER_ = 0.05). FC estimates for clusters exhibiting significant FC with the seed region across baseline and follow-up scans were extracted for subsequent data analysis. An exploratory networks analysis was conducted, using FC estimates within and between seven networks (default mode, ventral attention, dorsal attention, fronto-parietal control, visual, limbic, and somatomotor), which were delineated using Schaefer’s 400-region parcellation of the Yeo atlas.^30,31^

## Statistical analysis

### Clinical efficacy evaluation

The primary outcome was change in CH attack frequency per week (baseline vs. follow-up). In the event of multiple headache diary data entries per date, the higher CH attack entry was used. The secondary outcome was change in average pain intensity (baseline vs. follow-up). Statistical analyses for baseline vs. follow-up were conducted using the non-parametric Wilcoxon-signed-rank test. The family-wise error rate (FWER) for the two main clinical outcomes was controlled using the Bonferroni-Holm method (two tests, alpha: P_FWER_ < 0.05).^32^

### Functional and structural neuroimaging analysis

Change in hypothalamic FC was evaluated using Wilcoxon-signed-rank test. Simple linear regression was used to analyze the correlation between percent change in attack frequency from baseline to follow-up with change in FC. The FWER was controlled using the Bonferroni-Holm method (four tests).^32^

Exploratory analyses of psilocybin effects on brain structure (cortical thickness and GM volume) and on FC within and between networks were performed, evaluating change from baseline to follow-up and the correlation between percent change in attack frequency and change in neuroimaging outcome. Effect sizes (Cohen’s d and Pearson’s R) are reported as outcome measures, and due to the exploratory nature of these tests and to limit the number of statistical tests performed, we abstained from drawing statistical inference based on P-values for these analyses.

### Plasma psilocin analysis

We also explored a possible dose-response relation of maximum measured PPL (C_max_) and psilocin area under curve (AUC) with percent change in attack frequency and pain intensity, using linear regression (n=9, given incomplete psilocin data for Patient 2). Plasma psilocin AUC was calculated using the trapezoidal rule.

## References

1. Burish, M. J., Pearson, S. M., Shapiro, R. E., Zhang, W. & Schor, L. I. Cluster headache is one of the most intensely painful human conditions: Results from the International Cluster Headache Questionnaire. Headache 61, 117–124 (2021).

2. Headache Classification Committee of the International Headache Society (IHS). ICHD-3 beta. Cephalagia 33, 629–808 (2013).

3. Rozen, T. D. & Fishman, R. S. Cluster Headache in the United States of America: Demographics, Clinical Characteristics, Triggers, Suicidality, and Personal Burden. Headache 52, 99–113 (2012).

4. Schindler, E. a D. et al. Indoleamine Hallucinogens in Cluster Headache: Results of the Clusterbusters Medication Use Survey. J. Psychoactive Drugs 47, 372–381 (2015).

5. Sewell, R. A., Halpern, J. H. & Pope Jr., H. G. Response of cluster headache to psilcybin and LSD. Neurology 66, 1920–1922 (2006).

6. Karst, M., Halpern, J. H., Bernateck, M. & Passie, T. The non-hallucinogen 2-bromo-lysergic acid diethylamide as preventative treatment for cluster headache: an open, non-randomized case series. Cephalalgia 30, 1140–1144 (2010).

7. Morelli, N. et al. Functional magnetic resonance imaging in episodic cluster headache. J. Headache Pain 10, 11–14 (2009).

8. May, A. et al. Correlation between structural and functional changes in brain in an idiopathic headache syndrome. Nat. Med. 5, 836–838 (1999).

9. May, A., Bahra, A., Büchel, C., Frackowiak, R. S. J. & Goadsby, P. J. Hypothalamic activation in cluster headache attacks. Lancet (London, England) 352, 275–278 (1998).

10. Sprenger, T. et al. Specific hypothalamic activation during a spontaneous cluster headache attack. Neurology 62, 516–517 (2004).

11. Kraehenmann, R. et al. Psilocybin-Induced Decrease in Amygdala Reactivity Correlates with Enhanced Positive Mood in Healthy Volunteers. Biol Psychiatry 78, 572–581 (2014).

12. May, A., Bahra, A., Büchel, C., Frackowiak, R. S. J. & Goadsby, P. J. PET and MRA findings in cluster headache and MRA in experimental pain. Neurology 55, 1328–1335 (2000).

13. Madsen, M. K. et al. Psilocybin-induced changes in brain network integrity and segregation correlate with plasma psilocin level and psychedelic experience. Eur. Neuropsychopharmacol. 50, 121–132 (2021).

14. Ly, C. et al. Psychedelics Promote Structural and Functional Neural Plasticity. Cell Rep. 23, 3170–3182 (2018).

15. Raval, N. R. et al. A Single Dose of Psilocybin Increases Synaptic Density and Decreases 5-HT 2A Receptor Density in the Pig Brain. Int. J. Mol. Sci. 22, 1–14 (2021).

16. de Coo, I. F. et al. Increased use of illicit drugs in a Dutch cluster headache population. Cephalalgia 39, 626–634 (2019).

17. Di Lorenzo, C. et al. The use of illicit drugs as self-medication in the treatment of cluster headache: Results from an Italian online survey. Cephalalgia 36, 194–198 (2016).

18. Madsen, M. K. et al. Psychedelic effects of psilocybin correlate with serotonin 2A receptor occupancy and plasma psilocin levels. Neuropsychopharmacology 44, 1328–1334 (2019).

19. Nichols, D. E., Johnson, M. W. & Nichols, C. D. Psychedelics as Medicines: An Emerging New Paradigm. Clin. Pharmacol. Ther. 101, 209–219 (2017).

20. Madsen, M. K. et al. A single psilocybin dose is associated with long-term increased mindfulness, preceded by a proportional change in neocortical 5-HT2A receptor binding. Eur. Neuropsychopharmacol. 33, 71–80 (2020).

21. Nutt, D. J., King, L. A. & Phillips, L. D. Drug harms in the UK: a multicriteria decision analysis. Lancet 376, 1558–1565 (2010).

22. Amsterdam, J. van, Opperhuizen, A. & Brink, W. van den. Harm potential of magic mushroom use: A review. Regul. Toxicol. Pharmacol. 59, 423–429 (2011).

23. Sheehan, D. V et al. The Mini-International Neuropsychiatric Interview (M.I.N.I.): the development and validation of a structured diagnostic psychiatric interview for DSM-IV and ICD-10. J. Clin. Psychiatry 59 Suppl 20, 22–33;quiz 34-57 (1998).

24. Sewell, A. & Halpern, J. H. Psychedelic Medicine. in (eds. Winkelman, M. & Roberts, T.) 1–26 (Praeger Security International, 2007).

25. Studerus, E., Gamma, A. & Vollenweider, F. X. Psychometric evaluation of the altered states of consciousness rating scale (OAV). PLoS One 5, e12412 (2010).

26. Dittrich, A., Lamparter, D. & Maurer, M. 5D-ABZ: Fragebogen zur Erfassung Aussergewöhnlicher Bewusstseinszustände. Eine kurze Einführung. (PSIN Plus Publications, 2006).

27. Desikan, R. S. et al. An automated labeling system for subdividing the human cerebral cortex on MRI scans into gyral based regions of interest. Neuroimage 31, 968–980 (2006).

28. Behzadi, Y., Restom, K., Liau, J. & Liu, T. T. A component based noise correction method (CompCor) for BOLD and perfusion based fMRI. Neuroimage 37, 90–101 (2007).

29. Baroncini, M. et al. MRI atlas of the human hypothalamus. Neuroimage 59, 168–180 (2012).

30. Yeo, B. T. et al. The organization of the human cerebral cortex estimated by intrinsic functional connectivity. J. Neurophysiol. 106, 1125–1165 (2011).

31. Schaefer, A. et al. Local-Global Parcellation of the Human Cerebral Cortex from Intrinsic Functional Connectivity MRI. Cereb. Cortex 28, 3095–3114 (2018).

32. Holm, S. A Simple Sequentially Rejective Multiple Test Procedure. Scand. J. Stat. 6, 65–70 (1979).

