## Supplementary Information for "Psilocybin-induced reduction in chronic cluster headache attack frequency correlates with changes in hypothalamic functional connectivity"

| **Patient** | **Verbatim Comment** |
| --- | --- |
| 1 | Good experiences all three. Each time I had been there, it seemed the effect waned. Not something I feel like going to do for regular treatment. I didn't like the loss of control and therefore don't want to do it again. I didn't feel bad after, no bad after effects. I can relive the experience by listening to music, see colors for my inner eye. Wouldn't take it at home, but would like to take it under controlled conditions. |
| 2 | I had two good experiences and one experience that was sort of scary. But I felt well taken care of. But the experiences wouldn't deter me from doing it again. I guess that it is a treatment that would have to be repeated. If it were possible, I would try it again. |
| 3 | I had good experiences. I would be willing to trying it again. |
| 4 | I had no problems and felt completely safe. However, I can imagine that for some people it would not be something to do, like if you had had a psychosis. |
| 5 | I felt very safe. Nothing negative to say. I would recommend anyone with CH to do it because of the psychological benefits. Right now I'm going for a walk in the woods and enjoying it even though I just had 2 days of living hell due to cluster headache attacks. This is a substantial change for me, it has improved my life quality. |
| 6 | I felt safe, well taken care of. I had no subjective effects. I would like to participate again with a higher dose. |
| 7 | NA |
| 8 | It was a huge experience, my body was relaxed. I am more relaxed now during attacks than before. It was a safe experience all the way through. I think this could be taken further as a treatment. |
| 9 | Good experience, it's probably important not to be afraid of taking the drug. I think I had a partial effect on my cluster headache. Probably a bit anxiety-provoking if you haven't tried it before. I feel I learned some things about myself. The three times I took psilocybin, the experiences were very different. The last one I was very much out of my body. |
| 10 | I found it safe and very pleasant. I would like to try it every week. |

**SI Table 1.** Patient comments about their experience with psilocybin as a treatment.

|  | **Grey matter volume (Cohen’s d)** | **Cortical thickness (Cohen’s d)** | **Grey matter volume (Pearson's R)** | **Cortical thickness (Pearson's R)** |
| --- | --- | --- | --- | --- |
| Total grey matter | 0.03 | NA | -0.09 | NA |
| Cortical grey matter | 0.01 | -0.09 | -0.05 | 0.325 |
| Hippocampus | 0.05 | NA | 0.33 | NA |
| Lateral orbitofrontal | -0.39 | -0.84 | 0.65 | 0.077 |
| Medial orbitofrontal | -0.3 | -0.76 | 0.06 | -0.53 |
| Rostral middle frontal | -0.51 | -0.59 | -0.15 | -0.59 |
| Rostral anterior cingulate | -0.51 | -0.72 | 0.72 | 0.18 |
| Posterior cingulate | 0.14 | -1.11 | 0.28 | -0.03 |
| Caudal anterior cingulate | -0.13 | -0.44 | 0.51 | -0.12 |
| Pars orbitalis | -0.82 | -0.2 | 0.65 | -0.53 |
| Superior frontal | 0.21 | -0.54 | 0.42 | -0.42 |
| Supramarginal | -0.3 | -0.97 | -0.46 | 0.1 |
| Insula | 0.16 | -0.25 | 0.16 | 0.07 |
| Inferior parietal | 0.48 | -0.2 | 0.15 | -0.66 |
| Superior parietal | -0.62 | -1.62 | -0.08 | -0.36 |
| Inferior temporal | 0.15 | 0.04 | 0.09 | -0.05 |
| Middle temporal | 0.36 | -0.66 | 0.02 | -0.21 |
| Parahippocampal | 0.12 | -0.05 | 0.20 | -0.20 |

**SI Table 2. Effects of psilocybin on grey matter volume and cortical thickness.** Cohen’s d values as a measure of effect size (baseline vs follow-up), Pearson’s R from linear regression analysis of association between attack frequency percent change and structural change from baseline to follow-up (grey matter volume or cortical thickness).

| **Cluster** | **Peak voxel MNI coordinate** | **Size (voxels)** | **Size (mm^3^)** |
| --- | --- | --- | --- |
| **1. Diencephalon: hypothalamus, thalamus, caudate, brain stem** | +2,-10,-10 | 3294 | 26352 |
| **2. Left lingual gyrus** | -18, -44, 0 | 339 | 2712 |
| **3. Right cerebellum** | 14, -68, -12 | 221 | 1768 |
| **4 Occipito-temporal white matter** | 36, -30, 26 | 158 | 1264 |

**SI Table 3. Clusters exhibiting significant functional connectivity with hypothalamus seed.** MNI: Montreal Neurological Institute.

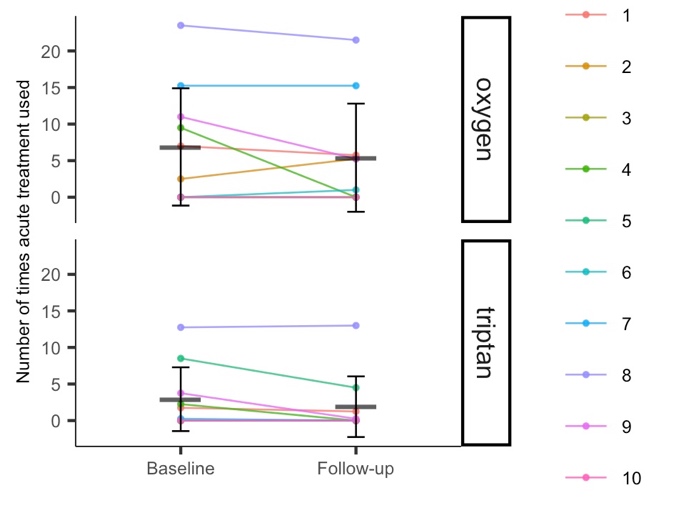

**SI Fig 1. Reported use of acute treatments.**

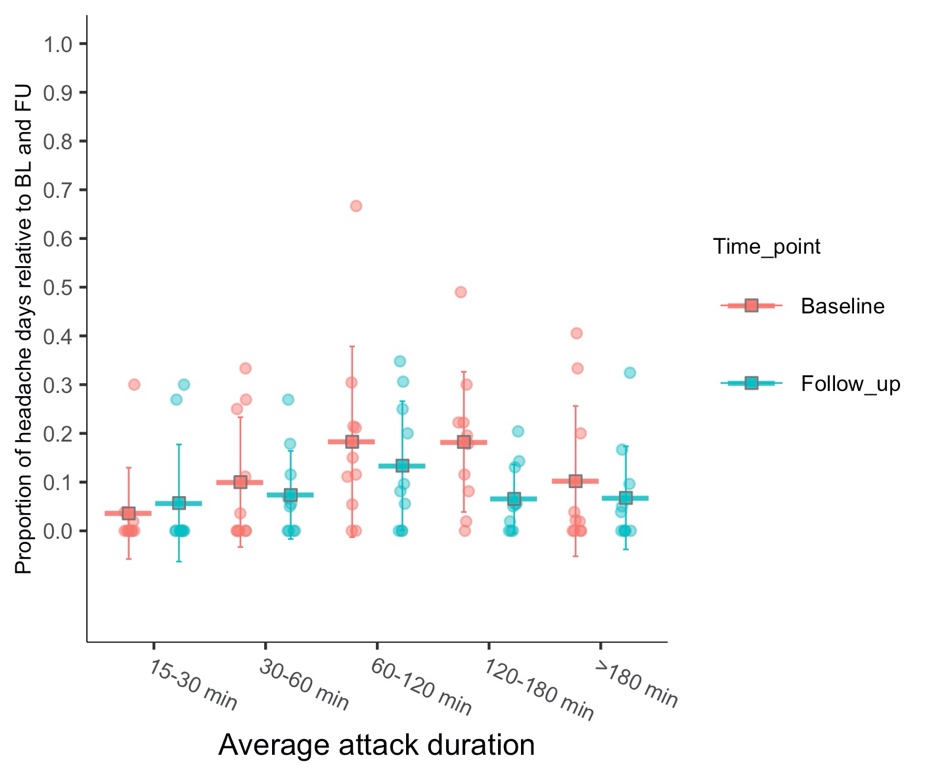

**SI Fig 2. Attack duration.** Proportion of average duration of attacks per day relative to baseline or follow-up. Error bars show mean and standard deviation.

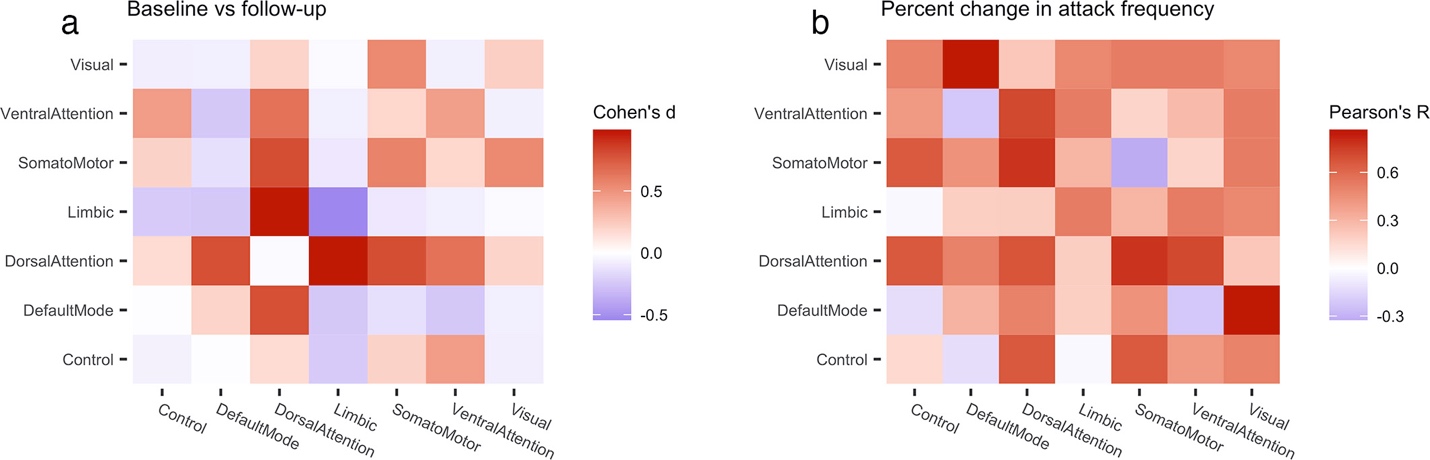

**SI Fig 3. Exploratory functional networks analysis.** a) Heat map shows Cohen’s d effect size estimates of change in functional connectivity from baseline to follow-up within and between functional networks. b) Heatmap showing correlation between percent change in attack frequency and change in functional connectivity from baseline to follow-up.
